# Aberrant systemic complement levels and altered acute-phase inflammatory responses attribute to varying grades of dengue disease severity

**DOI:** 10.1101/2025.09.04.25335083

**Authors:** Abdul R Anshad, Shanmugam Saravanan, Amudhan Murugesan, Vighnesh Ravindran, Sivadoss Raju, Rajendran Kannan, Yean K Yong, Marie Larsson, Esaki M Shankar

## Abstract

**Background:** Dengue virus (DENV) infection poses a serious health burden across the tropical and sub-tropical regions. Dengue manifestations range from asymptomatic and subclinical to severe disease with plasma leakage and organ dysfunction. The role of complement factors and acute-phase proteins in clinical dengue remains unclear.

**Methods:** Of the enrolled 156 participants, 114 were confirmed clinical dengue cases and 42 healthy controls. We performed serological profiling of NS1, anti-DENV IgM, and anti-DENV IgG, and measured serum acute-phase reactants, clinico-laboratory parameters, and viral load. These parameters were used for classification of disease severity in the patients namely, dengue with warning signs (DWS+, n=35), dengue without warning signs (DWS-, n=74), and severe dengue (SD, n=5) in accordance with the 2009 WHO guidelines. Levels of complement factors C1Inh, C1q, C2, C3a, C3b, MBL, C5a, and CR1 were assessed using commercial ELISA. The concentrations of these complement factors were correlated with various acute-phase proteins, clinical laboratory parameters, grades of dengue disease severity, and platelet counts.

**Results:** Serological classification revealed 104 patients were IgM positive, 35 were IgG positive, 24 were NS1 positive, and 26 were secondary dengue samples. There was a significant alteration in early classical complement pathway proteins C1Inh, C1q, and C2. The levels of downstream complement products and CR1 remained largely unchanged across both varying grades of dengue severity and primary/secondary stratification. MBL showed significant variation between the severity groups but did not differ within primary and secondary dengue samples. Univariate analysis revealed that NS1 positivity, IgG positivity, age, urea, and CR1 were factors correlated with the severity, but further multivariate analysis showed CR1 as the only independent predictor among complement factors that correlated negatively with dengue severity. Platelet counts had a negative association with RDW and basophils, and a strong positive correlation with uric acid levels.

**Conclusion:** Our findings demonstrate that aberrant complement activation contributes to varying grades of dengue severity. Moreover, CR1 may serve as a possible predictor of dengue severity.

**Author summary:** Dengue is a mosquito-borne tropical viral infection that can range in severity from asymptomatic to life-threatening manifestations. The human immune system represents a key determinant of dengue disease progression and severity. However, the specific mechanism that makes a subset of individuals in the population relatively severely ill is seldom understood. The complement system represents a key component of the natural immune system, which can help fend off infection but can also injures the host by exaggerating inflammation if not properly controlled. Here, we studied the role of certain important complement proteins and laboratory analytes in patients with dengue infection of varying grades of disease severity. We found that alteration of certain complement proteins i.e., C1Inh, C1q, and C2, depended on the severity of dengue infection. Notably, we found that the complement receptor 1 was inversely correlated with disease severity. We also observed associations between platelet counts and certain hematologic markers, including a strong positive correlation with uric acid. We concluded the role of aberrant complement activation, and identified CR1 (CD35) as a predictor of dengue disease severity.

## Introduction

Dengue virus (DENV) transmitted by female Aedes mosquitoes, represents an important global public health concern (1,2). Globally, an estimated 3.9 billion people are at risk of infection, mainly in the tropical and sub-tropical regions, with an eight-fold increase in incidence rates reported in the last two decades (3). Notwithstanding DENV infections are often asymptomatic or self-limiting, exacerbation could result in severe dengue disease (4). Four antigenically distinct serotypes, DENV1 - DENV4, appear to circulate in the population, while a fifth sylvatic serotype has also been identified among non-human primates (5,6). The disease spectrum and severity vary from unapparent sub-clinical infection to life-threatening severe dengue, which is often marked by severe thrombocytopenia and multi-organ failure (7). Progression of disease from mild to severe dengue is influenced by various factors like heterologous virus serotype and host inflammatory responses (8,9).

Of the various immunological and inflammatory pathways involved, the complement system represents an underexplored contributor of dengue pathogenesis. The complement system necessitates acute innate immune responses, and bridges the innate with the adaptive immune responses (10,11). Activation of the complement cascade, particularly via the alternative pathway, has been linked to increased levels of anaphylatoxins, driving the recruitment of polymorphonuclear neutrophils (PMNs) and NK cells, leading to excessive release of pro-inflammatory cytokines, and at times could contribute to endothelial dysfunction (12).

The complement system plays a critical role in dengue pathogenesis by inducing inflammation and tissue damage. The mannose-binding lectin (MBL) pathway appears to limit viral replication in asymptomatic and primary dengue disease. Elevated levels of complement-associated anaphylatoxins have been reported in severe secondary infection, whereas soluble immune complexes during acute dengue disease, will be bound by CR1 (CD35) expressed on erythrocytes following opsonization by complement factors (13). Despite these insights, there remains a lack of direct clinical data correlating complement protein levels with disease severity in dengue patients. Here, we quantitatively assessed the levels of key complement proteins using commercial ELISA, and several other key clinical laboratory parameters in dengue patients. We determined if complement components and acute-phase reactants correlated with disease severity attributing to improved understanding of dengue pathogenesis, and endeavoured to identify potential biomarkers/therapeutic targets in severe dengue.

## Materials and Methods

### Ethical approval

The cross-sectional case-control study was carried out in accordance with the Guidelines of the International Conference on Harmonization, and the Declaration of Helsinki (1964) to fulfil the study objectives. The Institutional Ethical Committee (IEC) of the Saveetha Medical College and Hospital, Chennai, (Ref. No. 114/03/2024/Faculty/SRB/SMCH), and the Government Theni Medical College and Hospital, Theni (Ref. No. 2300/IEC/2024-26) reviewed all the protocols and extended necessary approval for conducting the research. Written informed consents were duly obtained from all the study volunteers/patients. Peripheral blood specimens were collected from the recruited study participants in BD Vacutainer (Becton Dickinson, Franklin Lakes, NJ, USA) tubes for the extraction of serum by a trained phlebotomist. The schematic representation of the work flow involved in the investigations is illustrated in **Fig.1**.

**Figure 1.**
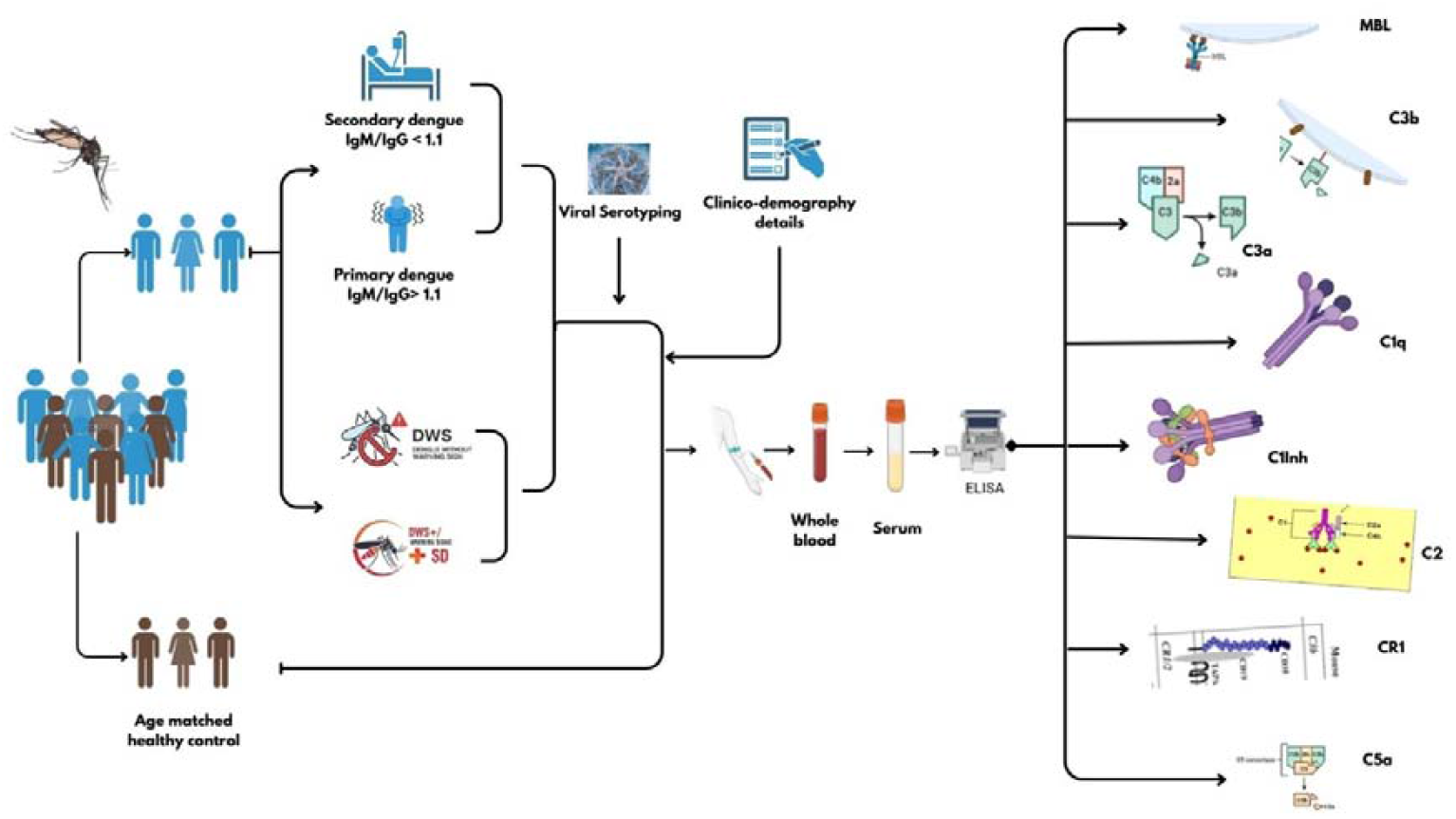
Graphical abstract of the cross-sectional case-control investigation (visual abstract created by Abdul R Anshad)

### Dengue diagnosis and clinical classification

This study encompassed patients diagnosed and admitted at the medical center with DENV infection, confirmed by laboratory methods such as RT-PCR or commercial ELISA-based detection of DENV-specific IgM antibodies or NS1 ELISA. Participants recruited at the clinical centres were classified as primary and secondary dengue infections. PanBio Dengue IgM (Cat. No.: 01PE20, Abbott, Chicago, IL, USA) and PanBio dengue IgG (Cat. No.: 01PE10, Abbott, Chicago, IL, USA) were employed for the characterization of IgM and IgG positive clinical samples. The samples were diluted 1: 100 in both assays as per the manufacturer’s instructions.

For the anti-DENV IgM assay, samples with a value >11 PanBio units were considered positive whereas samples with a value >22 PanBio units were considered positive for the anti-DENV IgG assay. Unless and otherwise mentioned, primary dengue infection was defined as those with an IgM/IgG ratio >1.2, and secondary dengue was defined as those with an IgM/IgG ratio higher or equal to 1.2 (14).

### Quantification of dengue viremia and serotyping

The serum samples were subjected to quantification of DENV viremia using commercial platforms. Briefly, RNA was extracted using a KingFisher^™^ Flex Purification system (ThermoFisher Scientific, Singapore) using HIPura Prefilled Medium Plates T (HiMedia, Mumbai, India). Subsequently, DENV viral load was quantified on a QuantStudio 5 Real Time PCR (Applied Biosystems, Foster City, CA, USA) using a commercial Dengue Real Time PCR kit (Cat. No.: 8013, Helini, Chennai, India) (15). The samples with confirmed viremia were used to identify the DENV serotype using a commercial DENV genotyping Real Time PCR kit (Cat. No.: 8014, Helini, Chennai, India).

### Chemiluminescence immunoassay (CLIA)

The presence of anti-SARS-CoV-2 IgG was measured in representative samples using a chemiluminescence immunoassay (CLIA). A total of 28 samples were selected randomly for the antibody measurements. As per the instructions of the manufacturer, the serum samples were pre-diluted with saline solution before measurement using IgG Quantitative reagent pack (Cat. No.: 6199960, Vitros immunodiagnostics, Bridgend, UK) in a CLIA system (VITROS 3600, Bridgend, UK).

### Complement ELISA

The products of complement cascade activation were measured by commercial ELISA. The following serum complement analytes were measured in the study participants as per the instructions of the manufacturers: mannose-binding lectin (MBL) (Cat. No.: DMBL00, Biotechne, Minneapolis, USA), C3b (Cat. No.: ab195431, Abcam), C3a (Cat. No.: ab279352, Abcam), C2 (Cat. No.: ab254501, Abcam), and C5a (Cat. No.: ab193695, Abcam), C1q (Cat. No.: ab170246, Abcam), C1 inhibitor (Cat. No.: ab224883, Abcam), and CR1/CD35 (Cat. No.: ab277439, Abcam).

### Statistical analysis

The comparison of demographic and clinical data was expressed as an interquartile range. The P values were calculated using the Chi-square test for categorical variables, while the continuous variables were analysed using the Kruskal-Wallis test. P value <0.05 was considered significant, and was used for the Post hoc Mann-Whitney U test. Univariate regression models were used to find significant relationships with outcome variables, which were subsequently included in the multivariate model. The Hosmer–Lemeshow values for the binary model and linear model were P=0.443 and 0.851, respectively. P values were *<0.05, **<0.01, and ****<0.0001. All the analyses were performed using PRISM Software ver.6.0 (GraphPad, California, USA) and SPSS 20.0 (IBM Corp, New York, USA).

## Results

### Clinico-demographic characteristics of the study cohort

The present study recruited 156 participants that encompassed DENV-infected patients (n=114; mean age 32 years) and healthy controls (n=42; mean age 27 years). Serological investigations (by commercial ELISA) revealed 92% participants (n=104) positive for anti-DENV IgM, 31% (n=35) positive for anti-DENV IgG, 21.2% (n=24) positive for DENV NS1, and 18 samples showed positivity for both DENV NS1 antigenemia and anti-DENV IgM levels. Based on the criteria discussed in the methodology, 26 patients were identified as secondary dengue infection for the study (**Table 1**). Of the 114 dengue positive cases, 47.4 % (n=74) patients were classified under dengue without warning signs, 22.4% (n=35) were categorized under dengue with warning signs, and 3.2% (n=5) were included under severe dengue.

**Table 1.** Cohort characteristics for demography, laboratory and serum complement analytes.

| Demographic and laboratory parameters | Total | Healthy controls | Dengue patients | P value |
| --- | --- | --- | --- | --- |
| Age, in years | 30 (23, 40) | 27 (22, 32.5) | 32 (25, 42) | 0.122 |
| Sex, male, *n* (%) | 111 (71.2) | 42 (100) | 69 (61.1) | 0.632 |
| Dengue category |  |  |  |  |
| DWS- | -- | -- | 74 (47.4%) | -- |
| DWS+ | -- | -- | 35 (22.4%) | -- |
| SD | -- | -- | 5 (3.2%) | -- |
| IgM positive, *n* (%) | -- | -- | 104 (92%) | -- |
| IgM titer | -- | -- | 27.9 (15.4, 57.8) | -- |
| IgG positive, *n* (%) | -- | -- | 35 (31%) | -- |
| IgG titer | -- | -- | 10.5 (4.9, 50) | -- |
| IgM/IgG ratio | -- | -- | 2 (0.9, 4.52) | -- |
| 2<sup>o</sup> infection, *n* (%) | -- | -- | 26 (23%) | -- |
| NS1 positive, *n* (%) | -- | -- | 24 (21.2%) | -- |
| C1Inh, pg/ml | 8.3 (7.78, 8.39) | 7.95 (7.5, 8.3) | 8.3 (8, 8.4) | 0.260 |
| C1q, ng/ml | 4.77 (4.4, 5.08) | 4.9 (4.75, 5.48) | 4.64 (4.3, 5.01) | 0.506 |
| C2, pg/ml | 8.08 (7.9, 8.35) | 8 (7.87, 8.16) | 8.11 (7.93, 8.43) | 0.774 |
| C3a, pg/ml | 8.4 (7.69, 8.73) | 8.45 (1, 8.78) | 8.35 (7.72, 8.73) | 0.297 |
| C3b, ng/ml | 4.52 (4.2, 4.81) | 4.7 (4.32, 5) | 4.45 (4.15, 4.75) | 0.946 |
| C5a, pg/ml | 5.36 (5.16, 5.49) | 5.37 (5.16, 6.49) | 5.36 (5.17, 5.5) | 0.566 |
| CR1, ng/ml | 0.47 (0.03, 0.72) | 0.48 (0.05, 0.8) | 0.47 (0.02, 0.69) | 0.096 |
| MBL, ng/ml | 3.04 (2.84, 3.36) | 3.21 (2.9, 3.49) | 3.02 (1.21, 4.58) | 0.781 |

**Footnotes:** All data are expressed as median (IQR) unless specified. *P* values are calculated by Chi square test for categorical variable and Kruskal–Wallis test for continuous variables. IQR, interquartile range; DWS-, Dengue without warning signs; DWS+, Dengue with warning signs; SD, Severe dengue; MBL, mannose-binding lectin

Viral load estimation was done for 80 samples, of which only nine samples were positive for dengue viremia. All the nine DENV viremia samples were concurrently positive for DENV NS1, three were positive for both NS1 and IgM and one was identified as secondary dengue. Similarly, of the 28 serum samples measured for SARS-CoV-2 IgG levels, all were observed to be reactive, with levels as high as 200 BAU/mL.

### Complement factors C1q, C1 inhibitor (C1 esterase) and C2 were differentially expressed and varied between grades of dengue severity

The analysis of complement components across the severity stages of DENV infection revealed that C1 Inh levels were significant higher for dengue patients without warning signs (P<0.001) and dengue with warning signs/severe dengue (P<0.01) compared to HCs. We also found higher levels in C2 between DWS- and HCs (P<0.5), and between DWS+/SD and HCs (P<0.05) The levels of C1q and MBL were significant lower between DWS- and HCs (P<0.01/P<0.05) and for DWS+/SD and HCs (P<0.01/P<0.05) and for **(Fig. 2)**. Conversely, activation products C3b, C3a, and C5a and the regulatory complement protein CR1 did not vary significantly across the dengue severity groups, which indicates downstream complement activation may not be uniformly altered during disease advancement.

**Figure 2.**
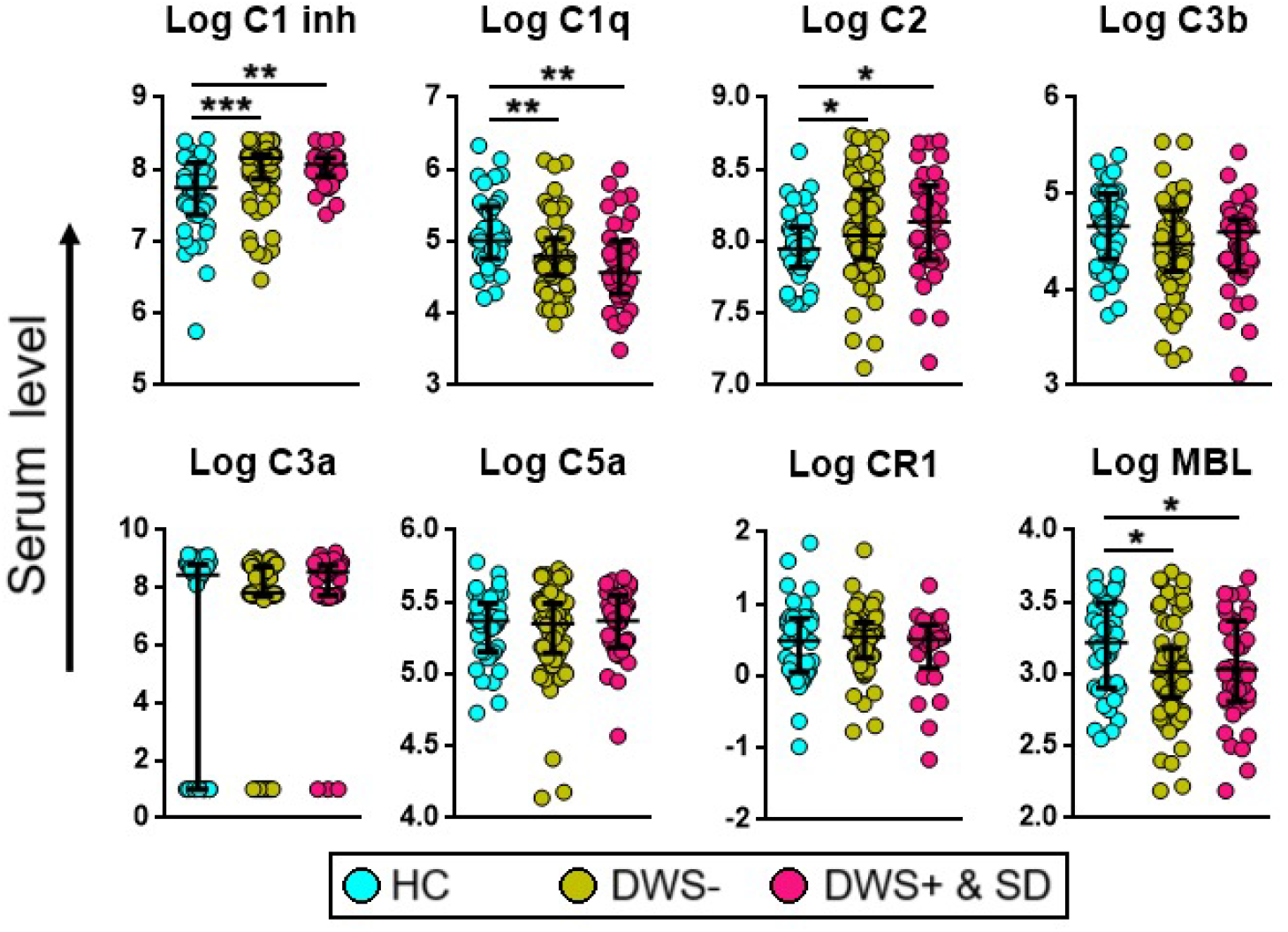
Comparison of serum levels of complement proteins C1 Inh, C1q, C2, C3b, C3a, C5a, CR1 and MBL in healthy controls, dengue without warning signs, as well as dengue with warning signs and severe dengue. Levels of biomarkers were compared across the three study groups by Kruskal–Wallis test. Post hoc Mann–Whitney U tests were subsequently performed for those complement biomarkers with a Kruskal–Wallis *P* value of <0.05. *P* value <0.05 (significant), where *<0.05, **<0.01, ***<0.001. **Footnotes:** HC, healthy controls; DWS-, dengue without warning signs; DWS+, dengue with warning signs; SD, severe dengue; C1Inh, C1 inhibitor; CR1, complement receptor-1; MBL, mannose-binding lectin

### Expression of C1 Inh, C1q, and C2 were significantly altered across primary and secondary dengue patients

Having said that the levels of C1q, C1 inhibitor and C2 are varied across grades of dengue severity, next we studied if their expressions varied between primary and secondary dengue patients. Analysis of complement factor levels among primary and secondary dengue patients, and healthy controls revealed a selective pattern. Early complement proteins including C1Inh levels were significantly higher in both primary (P=0.0003), and secondary dengue (P=0.0037) and HCs. There were significant lower C1q levels between primary dengue patients and HCs (P=0.0022), and between secondary dengue and HCs (P=0.0128). C2 levels were significantly lower in both primary (P=0.062), and secondary dengue (P=0.0005) and HCs, while activation products C3b, C3a, and C5a and regulatory protein, CR1 (P>0.5) did not vary significantly across the groups, which is in consistent with its expression in the severity. MBL did not show any significant variation between the groups unlike as in disease severity (**Fig. 3**).

**Figure 3.**
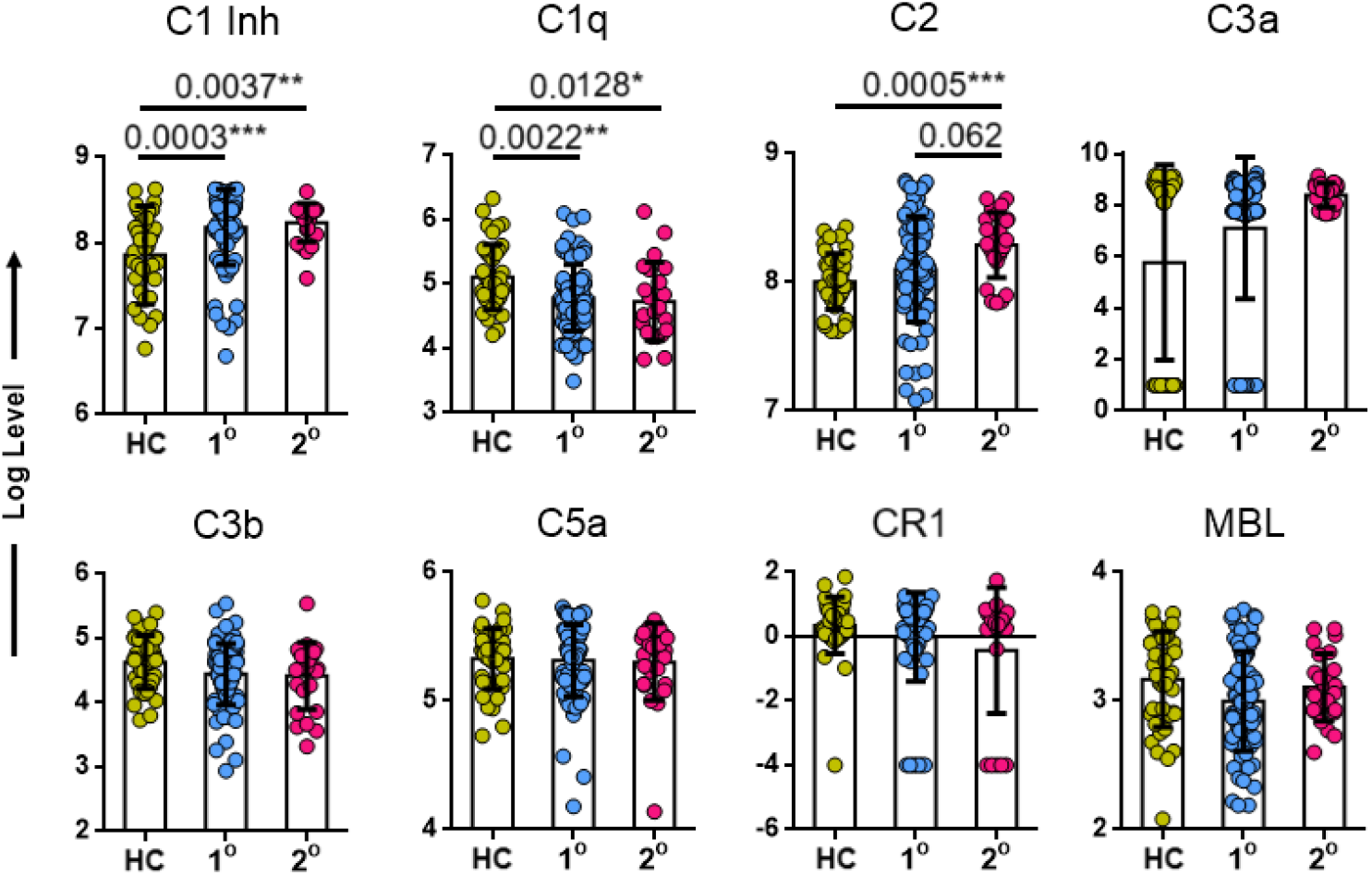
Comparison of serum levels of complement markers in healthy controls, 1^°^ – patients with primary dengue infection and 2^°^ – patients with secondary dengue infection. Levels of biomarkers were compared across the two patient groups and HCs by Kruskal– Wallis test. Post hoc Mann–Whitney U tests were subsequently performed for those biomarkers with a Kruskal–Wallis test P value of <0.05. P value <0.05 (significant), where *<0.05, **<0.01, ***<0.001. **Footnotes:** HC, healthy controls; DWS-, dengue without warning signs; DWS+, dengue with warning signs; SD, severe dengue; C1Inh, C1 inhibitor; CR1, complement receptor-1; MBL, mannose-binding lectin

### Every unit of increase in CR1 levels was significantly associated with reduced odds of dengue disease severity

Next, we used univariate regression models to unveil significant relationships with outcome variables, which were subsequently included in a multivariate model. Univariate analysis revealed that CR1, NS1 positivity, age, urea, and IgG positivity were significantly associated with dengue severity. However, in the multivariate logistic showed only CR1 showed a significantly association with disease severity, where with every unit increase of CR1 was associated with reduced odds of dengue severity by 22% (Coef.=0.776, 95% CI=0.588 – 0.925; P=0.046), suggesting a potential protective role (**Fig. 4A** and **4B**).

**Figure 4.**
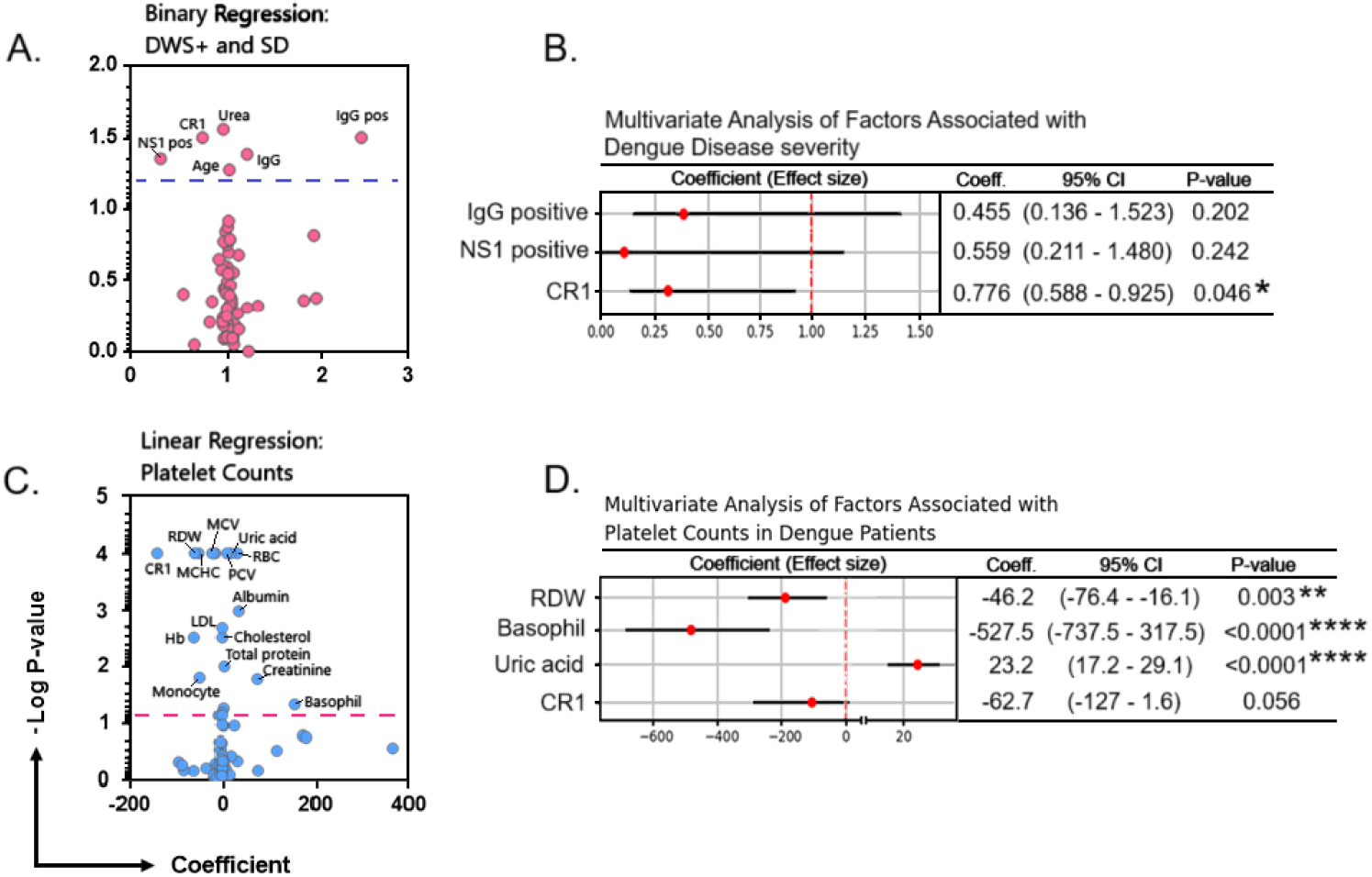
Factors that associated with dengue disease severity and platelet counts. **A)** Binary regression analysis of factors associated with severity of dengue disease (DWS+ and SD**). B)** Multivariate analysis of factors associated with dengue disease severity. **C)** Linear regression analysis of factors associated with platelet counts in dengue patients, and **D)** Multivariate analysis of factors associated with platelet counts in dengue patients. By using univariate regression model, variables that showed a significant relationship with outcome variables (viz. development of dengue with warning signs and or blood platelet counts) in univariate model will then be included in the multivariate model. The Hosmer–Lemeshow values for binary model and linear model were P=0.443 and P=0.851, respectively. *P* value <0.05 (significant), where *<0.05, **<0.01, and ****<0.0001.

### Serum uric acid levels were directly associated with blood platelet levels in clinical dengue infection

We assessed a wide range of hematologic and biochemical parameters/analytes for their relationship with blood platelet counts. Significant negative associations were observed with red cell distribution width (RDW) and peripheral basophil counts (P=0.003 and P<0.0001, respectively, whereas CR1 showed a negative trend but was not statistically significant (P=0.056). Concurrently, the levels of serum uric acid showed a strongly positive correlation with blood platelet counts (P<0.0001) (**Fig. 4C** and **4D**) by a multivariate analysis.

### Complement anaphylatoxins C3a and C5a showed significant positive correlations with immune markers and dengue viremia

Spearman correlation analysis revealed strong positive correlations with liver function markers ALT, AST, ALKP, GGT, and bilirubin with each other. On the other hand, lipid profile components, for instance LDL, HDL, TGL, VLDL, and cholesterol ratios showed both positive and negative correlations depending on the parameters. We also found correlation of certain acute phase inflammatory markers such as CRP, and ESR with neutrophils and lymphocytes. Certain renal function markers like creatinine, urea, and uric acid showed inter-correlations and some associations with plasma potassium and chloride electrolyte levels. We also found that the complement anaphylatoxins C3a, C5a, and C1q showing positive correlations with immune markers and possibly DENV viral load (**Fig. 5**).

**Figure 5.**
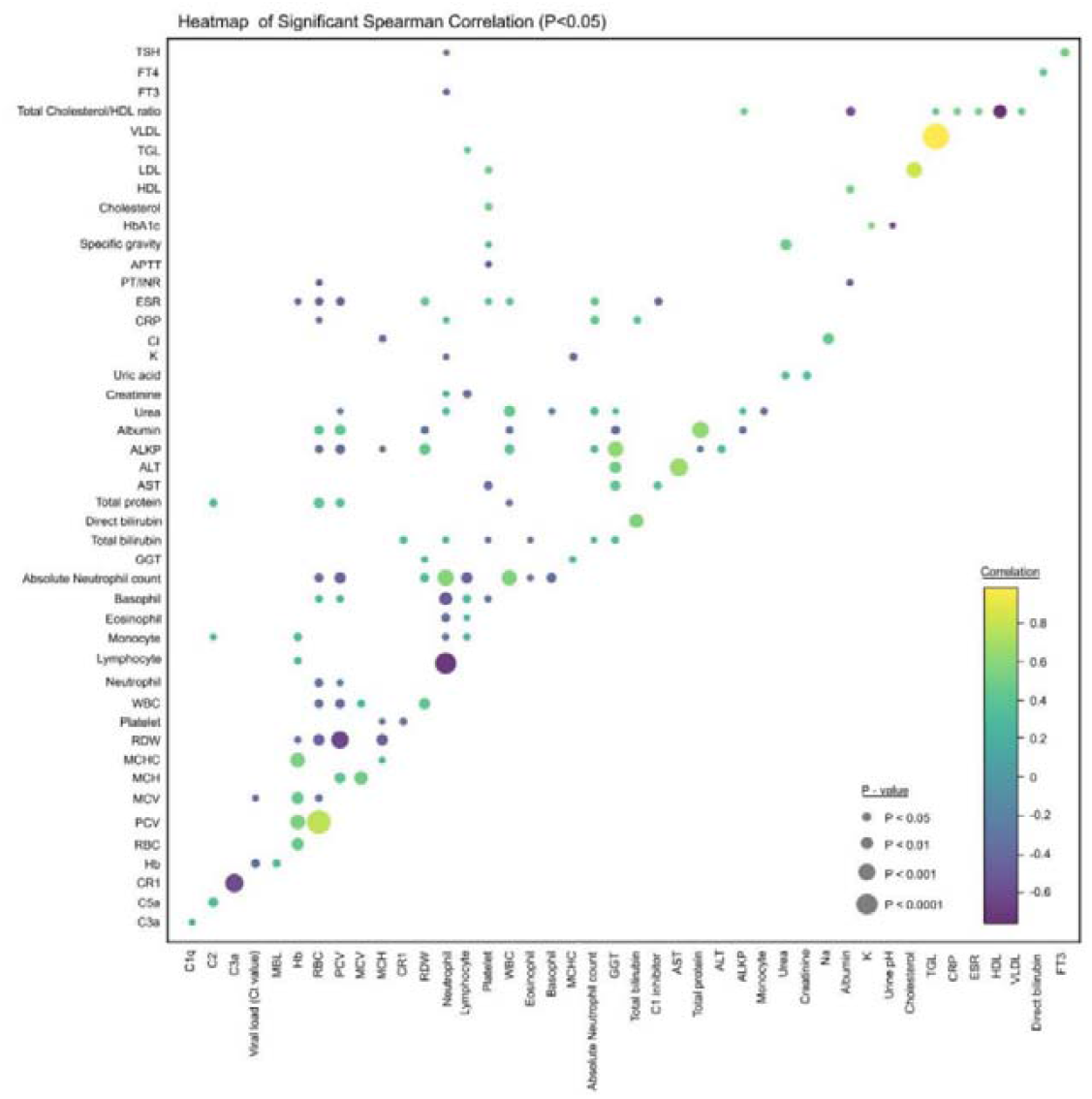
Heatmap of significant Spearman Correlations (P<0.05) among clinical, biochemical, hematologic, and immunological parameters. Warm color indicates positive correlation, dark color indicates negative correlation, and bubble size represents the level of significance.

## Discussion

In the current cross-sectional case control investigation, we surveyed the role of various clinical laboratory parameters and immunological determinants, with particular emphasis on complement system in DENV infected adults across varying grades of disease severity. Of the 156 study participant, 114 cases were serologically confirmed cases of DENV infection, with a predominance of IgM positivity and a small number of NS1 positive and secondary dengue cases. Viral RNA was also present in a minority of samples, all of which were NS1 positive, reinforcing the utility of NS1 as a marker of acute dengue viremia (16,17).

Analysis of complement components across stages of infection revealed distinct alterations in the early classical complement proteins. C1Inh (regulatory), C1q, and C2 demonstrated severity-dependent variations, suggesting that the classical pathway plays a crucial role in dengue immunopathogenesis. Our observations are consistent with prior evidence indicating the formation of an immune complex and antibody-mediated response can trigger the activation of classical pathway in dengue (18,19). Antibody-dependent enhancement (ADE) of dengue virus infection has been one of the main hypotheses to explain the disease severity of dengue (18). In presence of sub-neutralizing antibody, immune complexes can amplify the viral replication in phagocytic cells, which will in turn activate the complement system via cytokine cascade (9,20). The presence of C1Inh can be attributed to the compensatory mechanism by the host to counter excess activation of the complement proteins, thus limiting the inflammation and vascular mechanisms triggered by the same (21). Similarly, the attrition in C1q levels is consistent with its consumption during immune complex-driven activation, a well-defined hallmark of dengue disease, especially during secondary infection (18,22).

Our study also observed that MBL did not have any significant differential expression across primary and secondary dengue groups, although it did show a differential expression between varying grades of dengue disease severity. This observation, in turn, is consistent with the functional biology of MBL as a lectin complement pathway protein that recognizes carbohydrate moieties (17,23). Thus, the expression of MBL is not expected to vary remarkably between primary and secondary dengue, which primarily differentiates between humoral immune responses. On the contrary, the association of MBL with disease severity highlights its role in modulating host-virus interactions and complement activation during acute infection, making it more of marker associated with clinical outcome and not of disease severity (13,23). Our investigations also showed that the downstream complement activation products, viz., C3a, C3b, and C5a and the complement regulatory protein CR1 (CD35) did not show any significant difference in expression in varying grades of disease severity i.e. without warning signals, with warning signals and severe disease, as well as primary/secondary dengue. This suggests that terminal complement activation is not uniformly altered during disease severity, possibly due to efficient regulation or localized activation.

One of the most important observations was the association between CR1 and disease severity. By a univariate analysis, several parameters, including age, NS1 positivity, and IgG positivity, were found to be associated with dengue severity, while a multivariate modelling identified CR1 as the only factor associated with dengue disease severity. Furthermore, higher CR1 levels were protective, with each unit increase of CR1 reduces dengue severity by 22%. This suggests that CR1 exhibits a regulatory role in controlling complement activation, preventing excessive inflammation and tissue injury during systemic infection (18,24). Many previous studies have implicated uncontrollable complement activation as a contributor to plasma leakage, and our study hypothesizes the role of CR1 as a protective regulator (25). Also, CR1 showed a negative trend with platelet count but did not reach statistical significance, implicating that its suggestive role is more closely related to dengue severity than to thrombocytopenia.

Hematologic and biochemical results associated with the platelet count also provided additional insights. In line with previous findings (26,27), markers of immune derangement, such as RDW and basophil levels were inversely correlated with the platelet levels. These findings are consistent with the ongoing hematologic derangement occurring in dengue thrombocytopenia (26,27). Interestingly, uric acid levels showed a strong positive correlation with platelet count, raising the probability that altered purine metabolism or oxidative stress pathway can influence the hematologic recovery in DENV infection (28). However, this observation is only preliminary, and requires further in depth investigations.

The study has certain limitations that should be acknowledged. First, the relatively small sample size within the disease severity group might limit the statistical power and generalizability of the experimental findings. In addition, although we accessed complement activation, not all complement components were measured, which decreases the comprehensive understanding of the entire spectrum of the same. Furthermore, the cross-sectional design of the study limits the establishment of a temporal and causal relationship between the target analyte and disease exacerbation or mitigation over the period of disease progression warranting longitudinal follow-up investigations to capture the precise disease dynamics. Moreover, the possible interference of closely related flaviviruses that are (almost always subclinical, and indeed are representative confounding factors) circulating in our region should have been ruled out to claim that the aberrant responses observed herein are solely due to dengue. The study could have been strengthened significantly by including more diagnostic methods, such as a plaque reduction neutralization test to confirm DENV infection. Taken together, these limitations suggest this study should be considered as indicative, and a further large-scale, longitudinal study including more complement factors is warranted to validate the findings.

In conclusion, our investigation reinforces the importance of the complement system in dengue pathogenesis, with particular emphasis on the early classical pathway of complement activation. Also, the novel observation that CR1 acts as an independent protective factor against severity highlights its potential role as biomarker as well as therapeutic agent.

## Data Availability

All relevant data are within the manuscript and its Supporting Information files.

## Acknowledgements

The authors thank all the national and international members of the Infectious Diseases Society of India (http://www.idsi.org.in) (IDSI), Chennai, for extending insightful discussions as well as logistic support.

## Author contributions

A.R.A., and E.M.S., conceived and conducted the experiments. A.R.A., V.R., A.M., S.S., Y.K.Y., M.L., and E.M.S., designed the study and were responsible for conceptualization and data curation. A.R.A., M.L., and E.M.S., conducted the analysis, and were responsible for methodology, formal analysis, validation, and visualization. A.R.A, and E.M.S wrote the first draft of the manuscript. All authors reviewed and approved the manuscript.

## Funding

A.R.A. is funded by NFOBC-NBCFDC of the Ministry of Social Justice, Government of India Junior Research Fellowship. The study received funding support from M.L. through 201701091 the Swedish Research Council ("http://www.vr.se/english.html), Linköping University Hospital Research Fund, CALF, and the Swedish Society of Medicine. The funders of the study had no role in the study design, data collection, data analysis, data interpretation, or writing of the report.

## Conflict of interest

Authors does not have any conflict of interest

## Notes

### Competing Interest Statement

The authors have declared no competing interest.

### Funding Statement

Yes

### Author Declarations

Ethics committee/IRB of Saveetha Medical College and Hospital, Chennai, (Ref. No.114/03/2024/Faculty/SRB/SMCH), and the Government Theni Medical College and Hospital, Theni (Ref. No. 2300/IEC/2024-26) gave ethical approval for this work.

### Summary of Updates

Moreover, the possible interference of closely related flaviviruses that are (almost always subclinical, and indeed are representative confounding factors) circulating in our region should have been ruled out to claim that the aberrant responses observed herein are solely due to dengue. The study could have been strengthened significantly by including more diagnostic methods, such as a plaque reduction neutralization test to confirm DENV infection.

## References

1. Kurnia N, Kaitana Y, Salaki CL, Mandey LC, Tuda JSB, Tallei TE. Study of Dengue Virus Transovarial Transmission in Aedes spp. in Ternate City Using Streptavidin-Biotin-Peroxidase Complex Immunohistochemistry. Infect Dis Rep [Internet]. 2022 Sep 28;14(5):765–71. Available from: https://www.mdpi.com/2036-7449/14/5/78

2. Pan P, Zhang Q, Liu W, Wang W, Lao Z, Zhang W, et al. Dengue Virus M Protein Promotes NLRP3 Inflammasome Activation To Induce Vascular Leakage in Mice. Jung JU, editor. J Virol [Internet]. 2019 Nov;93(21). Available from: https://journals.asm.org/doi/10.1128/JVI.00996-19

3. Gowri Sankar S, Mowna Sundari T, Alwin Prem Anand A. Emergence of Dengue 4 as Dominant Serotype During 2017 Outbreak in South India and Associated Cytokine Expression Profile. Front Cell Infect Microbiol [Internet]. 2021 Aug 10;11. Available from: https://www.frontiersin.org/articles/10.3389/fcimb.2021.681937/full

4. Mukherjee S, Saha B, Tripathi A. Clinical significance of differential serum-signatures for early prediction of severe dengue among Eastern Indian patients. Clin Exp Immunol [Internet]. 2022 May 13;208(1):72–82. Available from: https://academic.oup.com/cei/article/208/1/72/6530528

5. Mustafa MS, Rasotgi V, Jain S, Gupta V. Discovery of fifth serotype of dengue virus (DENV-5): A new public health dilemma in dengue control. Med J Armed Forces India [Internet]. 2015 Jan;71(1):67–70. Available from: https://linkinghub.elsevier.com/retrieve/pii/S0377123714001725

6. Vaddadi K, Gandikota C, Jain PK, Prasad VSV., Venkataramana M. Co-circulation and co-infections of all dengue virus serotypes in Hyderabad, India 2014. Epidemiol Infect [Internet]. 2017 Sep 20;145(12):2563–74. Available from: https://www.cambridge.org/core/product/identifier/S0950268817001479/type/journal_article

7. Imad HA, Phumratanaprapin W, Phonrat B, Chotivanich K, Charunwatthana P, Muangnoicharoen S, et al. Cytokine Expression in Dengue Fever and Dengue Hemorrhagic Fever Patients with Bleeding and Severe Hepatitis. Am J Trop Med Hyg [Internet]. 2020 May 6;102(5):943–50. Available from: https://www.ajtmh.org/view/journals/tpmd/102/5/article-p943.xml

8. Mathew A, Rothman AL. Understanding the contribution of cellular immunity to dengue disease pathogenesis. Immunol Rev [Internet]. 2008 Oct 19;225(1):300–13. Available from: https://onlinelibrary.wiley.com/doi/10.1111/j.1600-065X.2008.00678.x

9. Nascimento EJM, Hottz ED, Garcia-Bates TM, Bozza F, Marques, Jr. ETA, Barratt-Boyes SM. Emerging Concepts in Dengue Pathogenesis: Interplay between Plasmablasts, Platelets, and Complement in Triggering Vasculopathy. Crit Rev Immunol [Internet]. 2014;34(3):227–40. Available from: http://www.dl.begellhouse.com/journals/2ff21abf44b19838,7f24896522e1c5c0,6593c20523f65461.html

10. Killick J, Morisse G, Sieger D, Astier AL. Complement as a regulator of adaptive immunity. Semin Immunopathol [Internet]. 2018 Jan 25;40(1):37–48. Available from: http://link.springer.com/10.1007/s00281-017-0644-y

11. Dunkelberger JR, Song WC. Complement and its role in innate and adaptive immune responses. Cell Res [Internet]. 2010 Jan 15;20(1):34–50. Available from: https://www.nature.com/articles/cr2009139

12. Carr JM, Cabezas-Falcon S, Dubowsky JG, Hulme-Jones J, Gordon DL. Dengue virus and the complement alternative pathway. FEBS Lett [Internet]. 2020 Aug 24;594(16):2543–55. Available from: https://febs.onlinelibrary.wiley.com/doi/10.1002/1873-3468.13730

13. Kraivong R, Punyadee N, Liszewski MK, Atkinson JP, Avirutnan P. Dengue and the Lectin Pathway of the Complement System. Viruses [Internet]. 2021 Jun 24;13(7):1219. Available from: https://www.mdpi.com/1999-4915/13/7/1219

14. Aggarwal C, Ahmed H, Sharma P, Reddy ES, Nayak K, Singla M, et al. Severe disease during both primary and secondary dengue virus infections in pediatric populations. Nat Med [Internet]. 2024 Mar 6;30(3):670–4. Available from: https://www.nature.com/articles/s41591-024-02798-x

15. Selvavinayagam ST, Sankar S, Yong YK, Anshad AR, Chandramathi S, Somasundaram A, et al. Serosurveillance of dengue infection and correlation with mosquito pools for dengue virus positivity during the COVID-19 pandemic in Tamil Nadu, India – A state-wide cross-sectional cluster randomized community-based study [Internet]. 2024. Available from: http://medrxiv.org/lookup/doi/10.1101/2024.06.07.24308595

16. de la Cruz-Hernández SI, Flores-Aguilar H, González-Mateos S, López-Martinez I, Alpuche-Aranda C, Ludert JE, et al. Determination of Viremia and Concentration of Circulating Nonstructural Protein 1 in Patients Infected with Dengue Virus in Mexico. Am Soc Trop Med Hyg [Internet]. 2013 Mar 6;88(3):446–54. Available from: https://www.ajtmh.org/view/journals/tpmd/88/3/article-p446.xml

17. Giang NT, van Tong H, Quyet D, Hoan NX, Nghia TH, Nam NM, et al. Complement protein levels and MBL2 polymorphisms are associated with dengue and disease severity. Sci Rep [Internet]. 2020 Sep 10;10(1):14923. Available from: https://www.nature.com/articles/s41598-020-71947-2

18. Diep NT, Giang NT, Diu NTT, Nam NM, Khanh L Van, Quang H Van, et al. Complement receptor type 1 and 2 (CR1 and CR2) gene polymorphisms and plasma protein levels are associated with the Dengue disease severity. Sci Rep [Internet]. 2023 Oct 13;13(1):17377. Available from: https://www.nature.com/articles/s41598-023-44512-w

19. Fuchs A, Lin TY, Beasley DW, Stover CM, Schwaeble WJ, Pierson TC, et al. Direct Complement Restriction of Flavivirus Infection Requires Glycan Recognition by Mannose-Binding Lectin. Cell Host Microbe [Internet]. 2010 Aug;8(2):186–95. Available from: https://linkinghub.elsevier.com/retrieve/pii/S1931312810002465

20. Byrne AB, Talarico LB. Role of the complement system in antibody-dependent enhancement of flavivirus infections. Int J Infect Dis [Internet]. 2021 Feb;103:404–11. Available from: https://linkinghub.elsevier.com/retrieve/pii/S1201971220325625

21. Yamanaka A, Hendrianto E, Mulyatno KC, Susilowati H, Ginting AP, Sary DD, et al. Correlation between Complement Component Levels and Disease Severity in Dengue Patients in Indonesia. Jpn J Infect Dis [Internet]. 2013;66(5):366–74. Available from: https://www.jstage.jst.go.jp/article/yoken/66/5/66_366/_article

22. Douradinha B, McBurney SP, Soares de Melo KM, Smith AP, Krishna NK, Barratt-Boyes SM, et al. C1q binding to dengue virus decreases levels of infection and inflammatory molecules transcription in THP-1 cells. Virus Res [Internet]. 2014 Jan;179:231–4. Available from: https://linkinghub.elsevier.com/retrieve/pii/S0168170213003973

23. Ornelas AM de M, Xavier-de-Carvalho C, Alvarado-Arnez LE, Ribeiro-Alves M, Rossi ÁD, Tanuri A, et al. Association between MBL2 haplotypes and dengue severity in children from Rio de Janeiro, Brazil. Mem Inst Oswaldo Cruz [Internet]. 2019;114. Available from: http://www.scielo.br/scielo.php?script=sci_arttext&pid=S0074-02762019000100326&tlng=en

24. Liu D, Niu ZX. The structure, genetic polymorphisms, expression and biological functions of complement receptor type 1 (CR1/CD35). Immunopharmacol Immunotoxicol [Internet]. 2009 Dec 30;31(4):524–35. Available from: http://www.tandfonline.com/doi/full/10.3109/08923970902845768

25. Schmidt CQ, Lambris JD, Ricklin D. Protection of host cells by complement regulators. Immunol Rev [Internet]. 2016 Nov 26;274(1):152–71. Available from: https://onlinelibrary.wiley.com/doi/10.1111/imr.12475

26. Castilho BM, Silva MT, Freitas ARR, Fulone I, Lopes LC. Factors associated with thrombocytopenia in patients with dengue fever: a retrospective cohort study. BMJ Open [Internet]. 2020 Sep 13;10(9):e035120. Available from: https://bmjopen.bmj.com/lookup/doi/10.1136/bmjopen-2019-035120

27. Ojha A, Nandi D, Batra H, Singhal R, Annarapu GK, Bhattacharyya S, et al. Platelet activation determines the severity of thrombocytopenia in dengue infection. Sci Rep [Internet]. 2017 Jan 31;7(1):41697. Available from: https://www.nature.com/articles/srep41697

28. Gandhi L, Maisnam D, Rathore D, Chauhan P, Bonagiri A, Venkataramana M. Differential localization of dengue virus protease affects cell homeostasis and triggers to thrombocytopenia. iScience [Internet]. 2023 Jul;26(7):107024. Available from: https://linkinghub.elsevier.com/retrieve/pii/S258900422301101X

